# A Mendelian randomization study identifies the causal association between plasma mitochondrial CHCHD proteins and polycystic ovary syndrome

**DOI:** 10.1101/2024.06.22.24309342

**Authors:** Shiyang Wei, Yafeng Wang, Niping Liu, Renfeng Zhao

## Abstract

**Purpose:** The objective of this research was to examine the causal link between PCOS and plasma mitochondrial coiled-coil-helix-coiled-coil-helix domain(CHCHD) proteins using a Mendelian randomization (MR) method.

**Methods:** We performed a two-sample MR analyses by utilizing summary statistics obtained from genome-wide association studies (GWAS) of PCOS (642 cases and 118,228 controls) and the levels of CHCHD2 and CHCHD10 in plasma (3,301 individuals). The inverse-variance weighted (IVW) method was used for the MR analyses, along with additional sensitivity analyses.

**Results:** The association between CHCHD2 and an increased risk of PCOS was identified (OR = 1.682; 95% CI = (1.231, 2.297); P = 0.001). The discovery of CHCHD10 revealed a protective impact on the likelihood of PCOS (OR = 0.828, 95% CI= 0.698-0.981, p = 0.029). The MR results were confirmed to be robust through the analysis of heterogeneity (P > 0.05) and pleiotropy (P > 0.05).

**Conclusion:** Our findings indicates that mitochondrial proteins CHCHD2 and CHCHD10 may play an important role in the pathogenesis of PCOS. Additional research is necessary to clarify the underlying mechanisms and investigate the potential of these proteins as targets for therapeutic intervention in PCOS.

**What does this study add to the clinical work:** A strong causal relationship has been established between two plasma mitochondrial complexes with coiled-coil-helix domains and polycystic ovary syndrome. The exact role of serum mitochondrial protein in polycystic ovary syndrome needs to be investigated via large-scale randomization trials or further studies.

## Introduction

Polycystic ovary syndrome (PCOS) is a prevalent hormonal condition that affects women during their childbearing years [1]. PCOS is characterized by hormonal imbalance, dysfunction of the ovaries, and the presence of numerous cysts in the ovaries [2]. PCOS has been linked to various metabolic disorders, including insulin insensitivity, obesity, and abnormal blood lipid levels [3]. The above-mentioned risk factors increase the likelihood of developing type 2 diabetes and cardiovascular disease [4,5]. The underlying mechanisms of PCOS remain unknown despite extensive research.

Mitochondria play an important role in cellular energy metabolism and physiological processes, including steroidogenesis, insulin signaling, oxidative stress response, apoptosis, and mitochondrial dynamics [6]. Studies suggested a possible relationship between mitochondrial dysfunction and PCOS [7,8]. Mitochondrial dysfunction in women with PCOS may contribute to metabolic abnormalities, such as insulin resistance and obesity [9,10]. Impaired mitochondrial function in PCOS may decrease energy production and increase oxidative stress, contributing to insulin resistance and hormonal imbalances [11]. Numerous studies have shown that mitochondrial enzymes involved in energy production are diminished in women with PCOS. However, the precise mechanisms by which PCOS is associated with mitochondrial dysfunction remain poorly understood. Moreover, genetic and environmental factors such as obesity and insulin resistance are hypothesized to play a role in developing mitochondrial dysfunction in PCOS [12,13].

Recent studies have established that mitochondrial coiled-coil-helix-coiled-coil-helix domain (CHCHD) proteins are involved in several cellular processes, such as mitochondrial function and response to oxidative stress [14,15]. However, the precise mitochondrial CHCHD proteins implicated in the development of PCOS and their causal connections to the disease are yet to be determined. Understanding the cause-and-effect relationship between these mitochondrial proteins and PCOS could provide substantial insights into the pathogenesis of the disease and potentially reveal novel therapeutic targets. Mendelian randomization (MR) is a statistical approach that uses genetic variants as instrumental variables (IVs) to investigate causal relationships between exposures and outcomes [16]. By minimizing biases from confounding and reverse causation, MR studies offer robust evidence for causal associations [17].

The present investigation employed MR analysis to ascertain the causal relationship between mitochondrial CHCHD protein concentrations and PCOS risk. We employed genetic variants as IVs for plasma protein levels to evaluate their effect on the risk of PCOS. Furthermore, we examined the reverse causation by analyzing how PCOS affects the plasma levels of mitochondrial CHCHD proteins. This two-way method allowed us to understand the causal correlation between mitochondrial CHCHD proteins and PCOS. The findings of this study could have substantial implications for the identification, prevention, and management of PCOS. Recognizing the causal link between mitochondrial CHCHD proteins and PCOS could aid the development of targeted therapies to restore mitochondrial function and improve PCOS-related metabolic abnormalities. Furthermore, determining the effect of PCOS on plasma concentrations of CHCHD proteins in mitochondria could help understand the possible role of mitochondrial dysfunction in the development of the syndrome.

## Methods

### Study design

To examine a causal relationship between PCOS and genetically predicted plasma levels of mitochondrial CHCHD proteins, we employed a two-sample MR strategy comprising two main stages. An MR analysis was initially performed using mitochondrial CHCHD proteins (CHCHD2 and CHCHD10) as exposures and PCOS as the outcome. In addition, an MR analysis was conducted in reverse, with PCOS as the independent variable and CHCHD2 and CHCHD10 as dependent variables. Notably, our study scrupulously maintains adherence to the following fundamental assumptions of MR analysis: (1) the independent variables used in the analysis strongly correlate with exposure, (2) the independent variables related to confounding factors have an insignificant influence on the relationship between exposure and the outcome, and (3) the independent variables solely affect the outcomes through their influence on exposure, excluding any other mechanisms. The study design is illustrated in Figure 1.

**Figure. 1.**
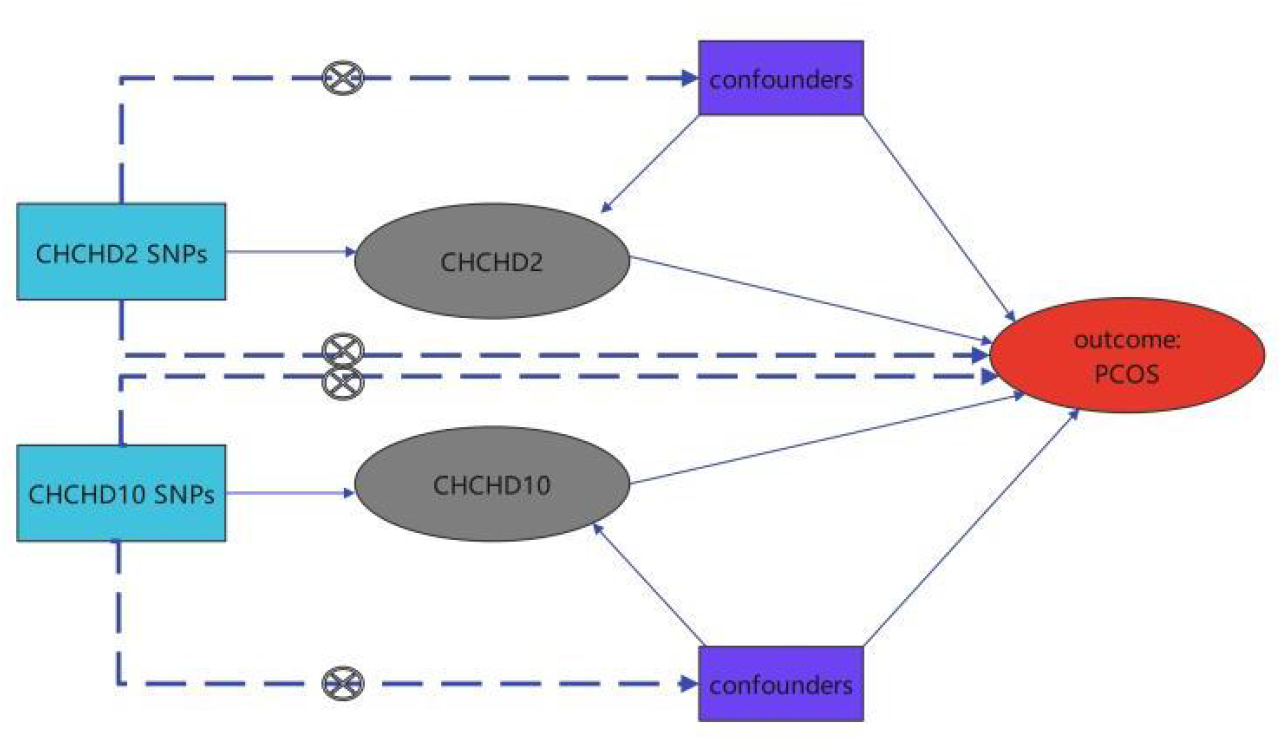
The flowchart of the MR study It is an illustrative diagram of the two-way MR analysis. We probed causalities by selecting important IVs for CHCHD2, CHCHD10, and PCOS. The flowchart demonstrates the three fundamental assumptions of the MR analysis.

### Data sources

GWAS summary data of CHCHD proteins and PCOS were obtained from the IEU OpenGWAS database (https://gwas.mrcieu.ac.uk/). The plasma mitochondrial CHCHD proteins (CHDCHD2 and CHCHD10) were acquired from a comprehensive GWAS on 3301 European ancestry healthy adults (ID ‘prot-a-534’ and ‘prot-a-535’). The genetic instruments for GWAS summary statistics of PCOS (642 cases and 118,228 controls) were retrieved from the FinnGen Consortium (ID ‘finn-b-E4_POCS’).

### Selection of the genetic instruments

Single nucleotide polymorphisms (SNPs) as IVs were utilized to assess the combined effect of mitochondrial CHCHD proteins on PCOS. We used p-values of 5×10^−6^ to select IVs for PCOS and plasma levels of mitochondrial CHCHD proteins. To ensure the selection of independent instruments, SNPs that did not overlap and were susceptible to LD clumping were chosen with a pairwise LD R2 threshold below 0.001. The genetic mutations were normalized by considering their impact while disregarding any palindromic mutations. The traits were analyzed by employing available genetic summary statistics for the variants. Moreover, to maintain the integrity of the results, we abstained from any modification or manipulation of the IVs, and the alleles linked to CHCHD2 and CHCHD10 were aligned in all the analyses. Instrument strength was determined by calculating the F-statistic, and IVs with F-statistics greater than 10 were considered robust.

### Statistical analysis

Inverse-variance weighted (IVW) was used as a key approach for univariate analysis in this study. A random-effects meta-analysis was performed to combine the results obtained from the individual SNPs. Additionally, weighted median, MR-Egger, weighted, and simple mode analyses were conducted to ensure the causal direction was consistent. Cochran’s test was performed in the IVW and MR-Egger models to assess heterogeneity among the included SNPs. Heterogeneity was considered significant if the p-value was < 0.05. Furthermore, we applied the MR-Egger regression to investigate potential pleiotropy, a method that allows for the simultaneous impact of genetic variants on multiple traits. In addition, MR-PRESSO detects and eliminates potential outliers that may introduce pleiotropy, consequently minimizing the probability of errors and bias due to pleiotropy. Statistical analyses were performed using R version 4.3.0.

## Results

### 3.1 Genetic IV selection for mitochondrial CHCHD proteins and PCOS

The current study used MR analysis to investigate the relationship between PCOS and CHCHD2 and CHCHD10. The research specifically examined the inverse correlation, with PCOS being identified as the risk and CHCHD proteins as the result. Based on the selection criteria, 12 and 10 SNPs were found to be associated with CHCHD2 and CHCHD10, respectively. To explore the reverse causal effect, eight SNPs were chosen as credible IVs representing PCOS as the exposure. Tables S1–3 contain detailed information on all the SNPs included.

### 3.2 The causal effect of CHCHD2 and CHCHD10 on PCOS

Figure 2 displays three sensitivity analyses (IVW, weighted median, and weighted mode) that confirmed the causal relationship between CHCHD10 and PCOS, whereas CHCHD10 and PCOS agreed with the two sensitivity analysis techniques (IVW and weighted median). This study discovered a notable link between CHCHD2 and CHCHD10 and the likelihood of PCOS, with corresponding odds ratio (OR) values of 1.682 (95% confidence interval [CI] 1.231–2.297; p = 0.001) and 0.828 (95% CI: 0.698–0.981; p = 0.001), respectively. A visual representation of the estimated effects of each genetic mutation on the outcomes can be observed in the forest plot (Figure 3). Figure S1 depicts a scatter plot depicting the estimated causal effects of each SNP on the risk of PCOS.

**Figure. 2.**
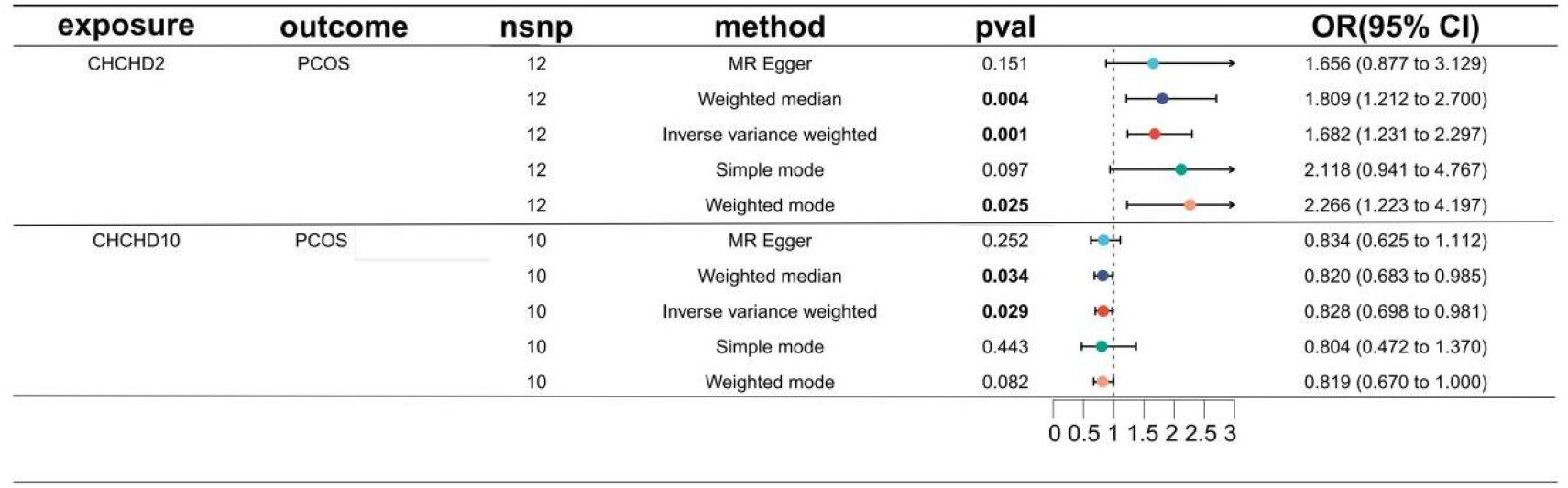
Results of MR analysis between CHCHD proteins and PCOS. Abbreviations: MR, Mendelian randomization; CI, confidence interval; OR, odds ratio; SNP, single nucleotide polymorphism; CHCHD, coiled-coil-helix-coiled-coil-helix domain-containing protein;

### 3.3 Sensitive analysis

Heterogeneity was assessed using the Cochran Q-test, which yielded p-values for MR-Egger and IVW. No significant differences were observed between the selected SNPs in CHCHD2 and CHCHD10 (Table 1). No indications of directional pleiotropy were detected in the MR-Egger regression. Figure S2 illustrates the results of the leave-one-out sensitivity analysis, revealing the causal connection between plasma CHCHD2, CHCHD10, and PCOS. The funnel plots of the associations between the SNPs in CHCHD2 and CHCHD10 and cancer risk were symmetrical, as presented in Figure S3.

**Table 1.**
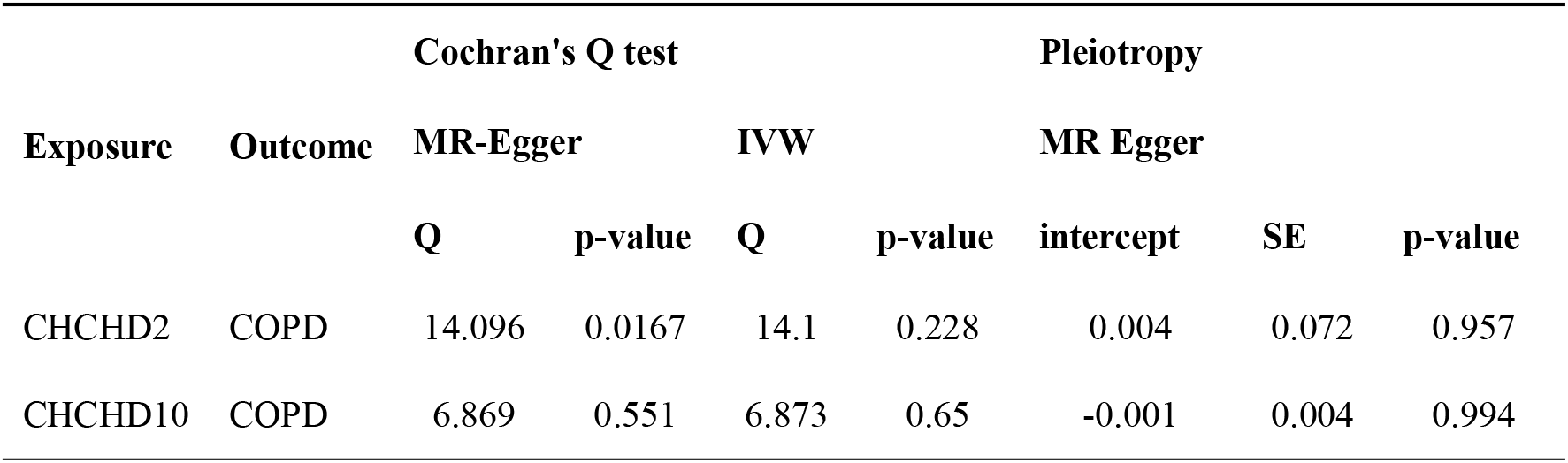
Results of sensitivity analyses of CHCHD proteins on PCOS. Abbreviations: MR, Mendelian randomization; IVW, inverse-variance weighted; CHCHD, coiled-coil-helix-coiled-coil-helix domain-containing protein; SE: standard error of the beta.

## Discussion

This study examined the causal link between plasma twin mitochondrial proteins, CHCHD2 and CHCHD10, and PCOS using MR analysis. The MR analysis evaluates causality by employing genetic variants as IVs. By employing genetic variations that have been established to be associated with exposure (CHCHD2 and CHCHD10) but not with outcome (PCOS), it is possible to reduce the probability of confounding bias and reverse causation. Furthermore, two-sample MR analysis demonstrated a noteworthy causal connection between the plasma proteins of the CHCHD domain and PCOS. We discovered a correlation between genetically anticipated elevated levels of CHCHD2 and heightened susceptibility to PCOS (OR = 1.682, 95% CI: 1.231–2.297, p = 0.001). In contrast, an inverse relationship was noted, suggesting that genetically projected higher plasma concentrations of CHCHD2 were linked to a reduced risk of PCOS (OR = 0.828, 95% CI: 0.698–0.981, p = 0.029). The current study found a potential causal association between CHCH2, CHCHD10, and PCOS development. Further research is required to elucidate the underlying mechanisms linking these proteins to PCOS.

Our findings exposed a notable and favorable connection between genetically anticipated plasma CHCHD2 concentrations and the risk of PCOS, while plasma CHCHD10 concentrations were negatively linked with PCOS risk. Mitochondrial respiration and oxidative phosphorylation are regulated by CHCHD2 [18]. Numerous neurodegenerative disorders, including amyotrophic lateral sclerosis (ALS) and Parkinson’s disease, have been associated with mutations in the CHCHD2 gene [19,20]. Moreover, CHCHD10 is involved in regulating mitochondrial structure and function [21]. Frontotemporal dementia, a rare neurodegenerative disorder, and cardiovascular diseases are linked to mutations in the CHCHD10 gene [22,23]. CHCHD2 and CHCHD10 genes play crucial roles in maintaining the health and functionality of mitochondria, which is essential for cellular metabolism [24,25]. ATP, an energy substrate, is synthesized via oxidative phosphorylation in mitochondria, earning them the moniker “powerhouses” of the cell [26]. Mitochondrial dysfunction can lead to various metabolic disorders, including mitochondrial diseases and neurodegenerative disorders [27]. Mitochondrial dysfunction may decrease ATP production and increase reactive oxygen species (ROS) generation, harming cells and tissues [28,29]. Several studies reported that women with PCOS have impaired mitochondrial function in various tissues, including the ovaries, skeletal muscle, and adipose tissues [30-32]. PCOS, insulin resistance, and abnormal steroidogenesis have been linked to mitochondrial dysfunction [33]. Ovaries of the PCOS mouse model exhibited reduced mitochondrial DNA content and altered gene expression related to mitochondrial biogenesis and function, impairing follicle development and, subsequently, oocyte quality [34,35]. Additionally, mitochondrial dysfunction in PCOS may contribute to developing insulin resistance [36]. Mitochondrial dysfunction can impede the pathways responsible for insulin signaling, leading to reduced glucose uptake and increased ROS production [37,38]. Consequently, insulin resistance, a common characteristic of PCOS, may occur [39]. The precise mechanisms by which mitochondrial dysfunction is associated with PCOS remain unknown. Our findings further supported that the involvement of CHCHD2 and CHCHD10 in mitochondrial dysfunction plays a potential role in the development of PCOS. The interplay between genetic and environmental factors likely contributes to mitochondrial dysfunction in PCOS [40]. Disruptions of mitochondrial function may also be influenced by hormonal imbalances, including increased androgen and insulin levels [41,42].

The findings of this study could have substantial implications for understanding the underlying mechanisms of PCOS and identifying potential therapeutic targets. Targeting mitochondrial dysfunction and oxidative stress pathways may offer novel therapeutic and preventive strategies for PCOS. Further research is required to elucidate the distinct functions of CHCHD2 and CHCHD10 in PCOS development and explore possible interventions that can regulate their levels or influence their activity. Future research should focus on the specific molecular mechanisms underlying the association between CHCHD2 and CHCHD10 levels and metabolic abnormalities in PCOS.

In conclusion, this study indicates a possible causal link between twin mitochondrial proteins, CHCHD2 and CHCHD10, in the plasma and PCOS using MR analysis. Our results suggested that mitochondrial dysfunction may contribute to PCOS development. Additional investigations are required to clarify the fundamental processes and investigate the healing possibilities of focusing on the levels of CHCHD2 and CHCHD10 for PCOS management.

### Limitations and Strengths

This study had some limitations. The MR analysis is predicated on the supposition that the genetic variants used as IVs are valid instruments and satisfy the requirements of instrumental variable assumptions. Despite utilizing previously validated genetic variants linked to CHCHD2 and CHCHD10 levels, the results may still be biased due to pleiotropy or horizontal pleiotropy. Notably, our research included individuals from a particular ethnic group, potentially restricting the applicability of our results to different demographics. As far as our knowledge goes, however, this is the first study to investigate the correlation between CHCHD proteins and PCOS. Future studies should verify these findings in diverse populations.

## Data Availability

All data produced in the present study are available upon reasonable request to the authors

## Acknowledgments

The authors express their gratitude for the valuable data resources provided by the researchers and databases.

## Ethics statement

As this was a reanalysis of published data, no ethical approval was required in this study.

